# The epidemic dynamics of three childhood infections and the impact of first vaccination in 18^th^ and 19^th^ century Finland

**DOI:** 10.1101/2022.10.30.22281707

**Authors:** Michael Briga, Tarmo Ketola, Virpi Lummaa

**Author notes:** Corresponding author: Michael Briga. Author Contributions: All authors contributed to the study design, the data collection and the writing of the manuscript. M.B. performed the analyses. Competing Interest Statement: The authors declare no competing interests.

## Abstract

Childhood infectious such as smallpox or measles have devasted human populations, but our knowledge on the history of public health interventions remains limited. Here, we use 100 years of newly available records in 18^th^ and 19^th^ century Finland to investigate the epidemic dynamics of three infections, smallpox, pertussis and measles and the impact of the country’s first nationwide vaccination programme. Between 1750 and 1850, we found over 40 epidemics of smallpox, pertussis and measles, which together were responsible for almost 20% of all registered deaths under age 10. The start of the first vaccination programme against smallpox in 1802 promptly triggered five major changes in smallpox epidemiology: (i) decreasing mortality, (ii) increasing age at infection, (iii) increasing time between epidemics, (iv) increasing fade-outs, which all result from (v) decreased pathogen transmission. In contrast, most epidemic characteristics of the childhood infections without vaccines, pertussis and measles, changed in the opposite direction to that of smallpox: their death toll almost doubled, their reproduction number increased and pertussis, but not measles, showed a decreasing age at infection and accelerating epidemics. Our study captures a rare glimpse of the epidemiology of childhood infections in historical populations and the possible long-term impact of major public health interventions.

## 1. Introduction

Infectious diseases have devasted human societies. Smallpox and measles decimated Europe and native American populations (1). While smallpox is relatively unique, with over 200 years of public health efforts having led to its eradication in 1980 (1), other childhood infections such as pertussis and measles persist or are even on the rise despite more than 50 years of intense vaccination efforts, a phenomenon attributed to, among others, vaccine hesitancy, transient epidemic dynamics such as the ‘honeymoon effect’ or pathogen evolution (2, 3).

Much can be learned from studying epidemic dynamics and public health interventions from historical societies (4–6). For example, mortality due to childhood infections is a driving force behind human reproductive rates and the associated demographic transition (7, 8). More recently, historical pandemics, such as the 1918 influenza pandemic, provided lessons for Covid-19 interventions (9) and studying historical vaccination laws can inform the optimization of vaccine implementation in the face of high vaccine hesitancy (10).

Studying infectious diseases in historical societies has proven challenging. Accurate historical data remain rare, often have a limited follow-up time or use annual records and linking these data to specific historical events remains difficult. In this study, we present a new database containing nation-wide daily records on births, deaths and causes of death in 18^th^ and 19^th^ century Finland, a pre-demographic transition population with high birth and childhood mortality rates (11, 12). We use this database for two purposes. First, we provide a detailed description of the historical epidemic dynamics of three childhood infections smallpox, pertussis and measles in a pre-demographic transition society. Second, Finland introduced annual vaccination against smallpox in 1802, with a strong bias towards children and a coverage starting from approximately 50% in 1812 and rising to 75% in 1850 (10, 13). We use the roll-out of this vaccination programme to test five theoretically expected impacts of vaccination: (i) decreasing mortality, (ii) increasing age at infection, (iii) increasing time between epidemics, (iv) increasing fade-outs, which all result from (v) decreased pathogen transmission (14, 15). Because we have data on more than one infection, we can compare the impact of vaccination against smallpox with that of two ‘control’ infections, pertussis and measles, that have similar transmission and epidemiology as smallpox, but for which public health interventions did not exist at that time.

## 2. Results

After data cleaning, selection and control procedures (see Methods section 4.1; Supplementary Information S1; Fig. S1), we obtained a database running from 1750 until 1850 containing 213 parishes (Fig. 1A) with 1,693,056 birth records and 1,190,627 death records. During this observation period, Finland’s population size increased fourfold from almost 400,000 to 1,600,000 inhabitants (Fig. 1B) and the data cover 50% of Finland’s population at any given time (interannual SD: 5%; Fig. S2). We provide more information on these data, their selection and their validation in the methods section and in the Supplementary Information Section 1.

**Fig. 1.**
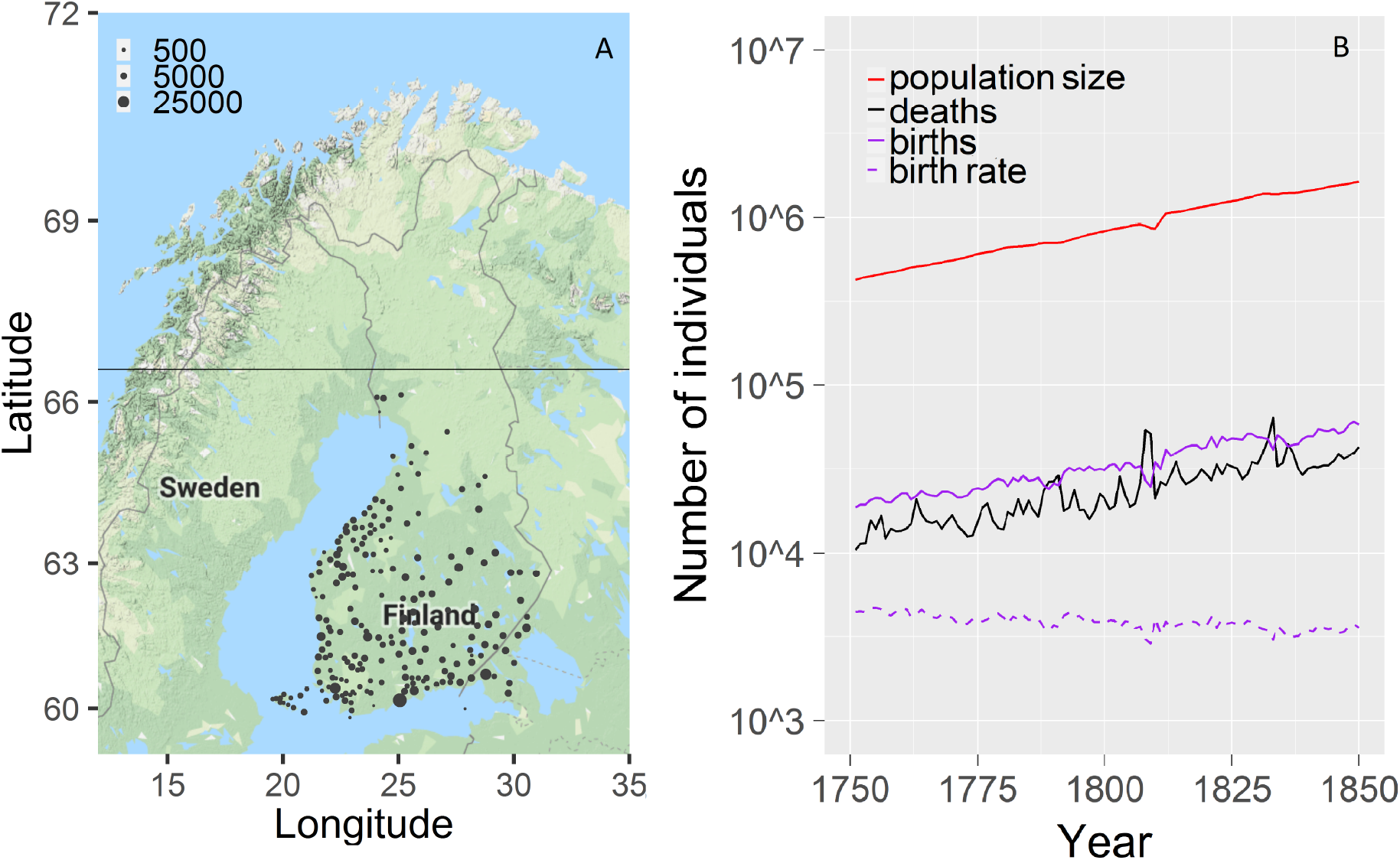
Data description with (A) geographic distribution, showing the location and number of inhabitants of 213 parishes located south of the Arctic Circle (horizontal line) and (B) the demography of the study population following the reports by the Official Statistics of Finland retrieved from https://www.stat.fi/index_en.html on November 1^st^ 2018. For graphical purposes, birth rate is expressed as the number of births per 100,000 population size.

In our database childhood mortality rates were high. Before the introduction of smallpox vaccine, i.e., between 1750 and 1801, further referred to as the *pre-vaccine era*, the database contained 694,192 registered births, of which 24% died by the age of 1 year, 37% by age 5 and 41% by age 10 (Table S1; Fig. S3). After the introduction of the smallpox vaccine, i.e., from 1802 until 1850 and further referred to as the *vaccine era*, our database contained 997,700 registered births and childhood mortality decreased with almost 2% compared to the pre-vaccine era, with 22% of all registered births dying under age 1, 35% under age 5 and 40% under age 10 (Table S1; Fig. S3). After age 10, deaths by the three infections remain rare (Fig. S4, results section 2.2)

### 2.1 Time series of infectious disease mortality

First, we investigated the contributions of childhood infections to childhood mortality in 17^th^ and 18^th^ century Finland. We focused on three readily identifiable childhood infections: smallpox, pertussis and measles (see Methods section 4.1). Between 1750 and 1850, we identified for each infection at least 15 epidemics (Fig. 2). The three infections played a prominent role in the high childhood mortality rate. In the pre-vaccine era, they were responsible for 11% of all registered deaths by age 1 year and this increased to almost 20% by the age of 5 or 10 years (Table S1; Fig. S4). In the vaccine era, the three infections were responsible for 12% of the deaths by age 1 year, but at older ages, their contributions decreased relative to the pre-vaccine era to 18% of the deaths by the age of 5 or 10 years (Table S1; Fig. S4).

**Fig. 2.**
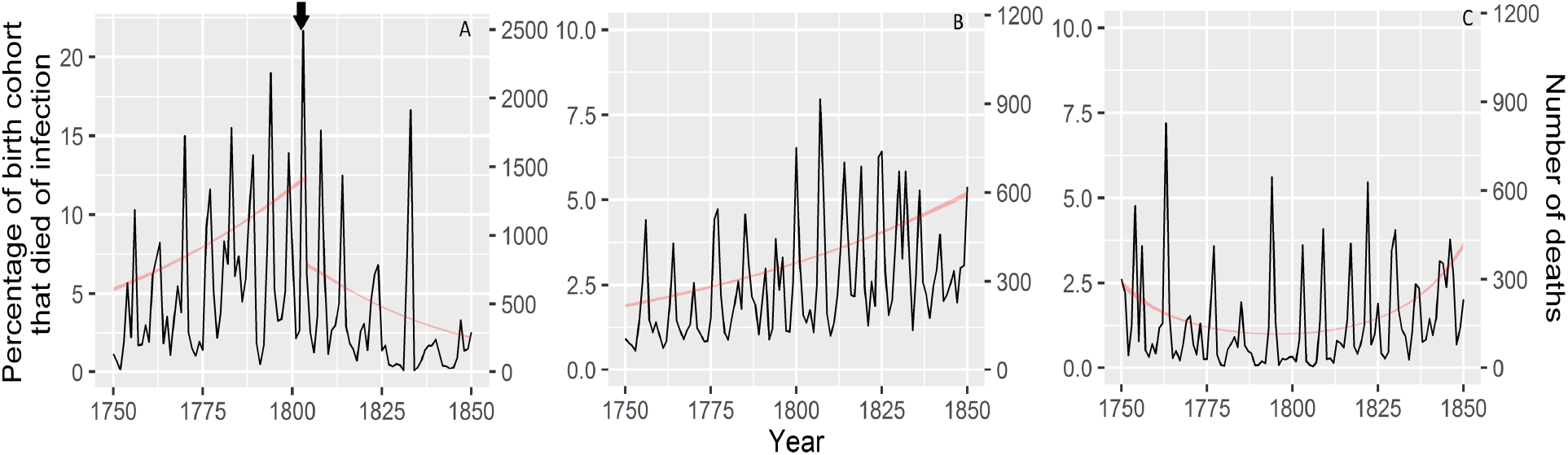
Over 40 recurrent epidemics of (A) smallpox, (B) pertussis and (C) measles between 1750 and 1850 in Finland. Vaccination immediately reduced smallpox mortality, while in the same period the mortality of childhood infections without vaccination, pertussis and measles, increased. Black arrow indicates the first record of a smallpox vaccine administered in Finland in 1802. Red line shows the best fitting model fitting the time series in infectious disease mortality (±95CI).

Second, we investigated the temporal trends in smallpox mortality and whether we could detect an effect of vaccination. Model selection indicated that there was a change in smallpox mortality, which occurred abruptly rather than gradual (threshold models fitted the data better than models with year as a continuous variable ΔAICc=-1.0; Fig. 2; Table S2A). The threshold model indicated that the change occurred in 1805 (95CI: 1804-1806), i.e., three years after the introduction of smallpox vaccination. Indeed, before 1805, smallpox mortality increased with time, while after 1805, smallpox mortality changed in the opposite direction (Fig. 2; Table S2A). GAM and its derivative identified a somewhat earlier turning point, namely in the year 1799 (95CI: 1798-1800). Thus, the rise in smallpox mortality broke quickly after or around the introduction of smallpox vaccination and was followed by a declining trend.

We then investigated the changes in the two ‘control’ infections pertussis and measles. In contrast to smallpox, for pertussis showed a linear or accelerating increase with time (−6.6<ΔAICc<-4.7; Fig. 2; Table S2B). For measles, the mortality declined until the early 1800’s after which it increased (ΔAICc=-6.8; Fig. 2; Table S2C). Thus, after vaccination, smallpox showed an abrupt decline in mortality, while at the same time the mortality from the infections without vaccination, pertussis and measles, increased.

### 2.2 Age at infection

The deaths due to three childhood infections were strongly biased towards children, with 95% of their deaths occurring in children under the age of 10 years (Table 1; Fig. 3; Fig. S5 A&B). In the pre-vaccine era, the mean age at death for pertussis was 2.3 years (95CI: 2.2-2.4), for measles it was 3.3 years (95CI: 3.2-3.4) and for smallpox it was 3.9 years (95CI: 3.8-3.9). The differences in the mean age at death between infections were strongly statistically supported (ΔAICc=-2497; Table S3A).

**Table 1.** Overview of the infections in this study with the descriptive statistics (±95CI) for each infection in the pre-vaccine (1750-1801) and vaccine eras (1802-1850). N = sample size; A_50_= mean age at death in years; A_95_ 95 percentile of the age at death distribution; fade-out = number of weeks per year without infectious death either nationwide or per parish.

| Infection | Smallpox |  | Pertussis |  | Measles |  |
| --- | --- | --- | --- | --- | --- | --- |
| pathogen | virus |  | bacteria |  | virus |  |
| generation time | 2 weeks |  | 4 weeks |  | 2 weeks |  |
| vaccination status | vaccine in 1802 |  | control (no vaccine) |  | control (no vaccine) |  |
| Predictions regarding the impact of vaccination on: |  |  |  |  |  |  |
| mortality | decrease |  | NA |  | NA |  |
| age at infection | increase |  | NA |  | NA |  |
| period | increase |  | NA |  | NA |  |
| fade-out | increase |  | NA |  | NA |  |
| R<sub>0</sub> | decrease |  | NA |  | NA |  |
| Data | 1750-1801 | 1802-1850 | 1750-1801 | 1802-1850 | 1750-1801 | 1802-1850 |
| vaccination | no | yes | no | no | no | no |
| N births | 694,192 | 997,700 | 694,192 | 997,700 | 694,192 | 997,700 |
| N deaths | 39,082 | 30,629 | 14,385 | 31,175 | 7,429 | 15,153 |
| N deaths per year | 752 | 638 | 277 | 649 | 143 | 316 |
| A<sub>50</sub> | 3.9 (3.8-3.9) | 4.7 (4.7-4.8) | 2.3 (2.2-2.4) | 1.8 (1.8-1.8) | 3.3 (3.2-3.4) | 3.4 (3.4-3.5) |
| A<sub>95</sub> | 11 (10.9-12.5) | 17.5 (17.0-18.2) | 6.5 (6.3-6.7) | 5.5 (5.2-5.7) | 9 (8.4-10.1) | 10.5 (10.4-10.6) |
| Nationwide fade-out | 3.0 (0-17.9) | 4.8 (0-34.0) | 4.3 (0-19.5) | 0.4 (0-2.8) | 18.1 (0.3-41.7) | 8.4 (0-37.6) |
| Parish-specific fade-out | 48.5 (25-53) | 50.4 (35-53) | 50.0 (35-53) | 49.7 (35-53) | 50.9 (39-53) | 51.0 (42-53) |

**Fig. 3.**
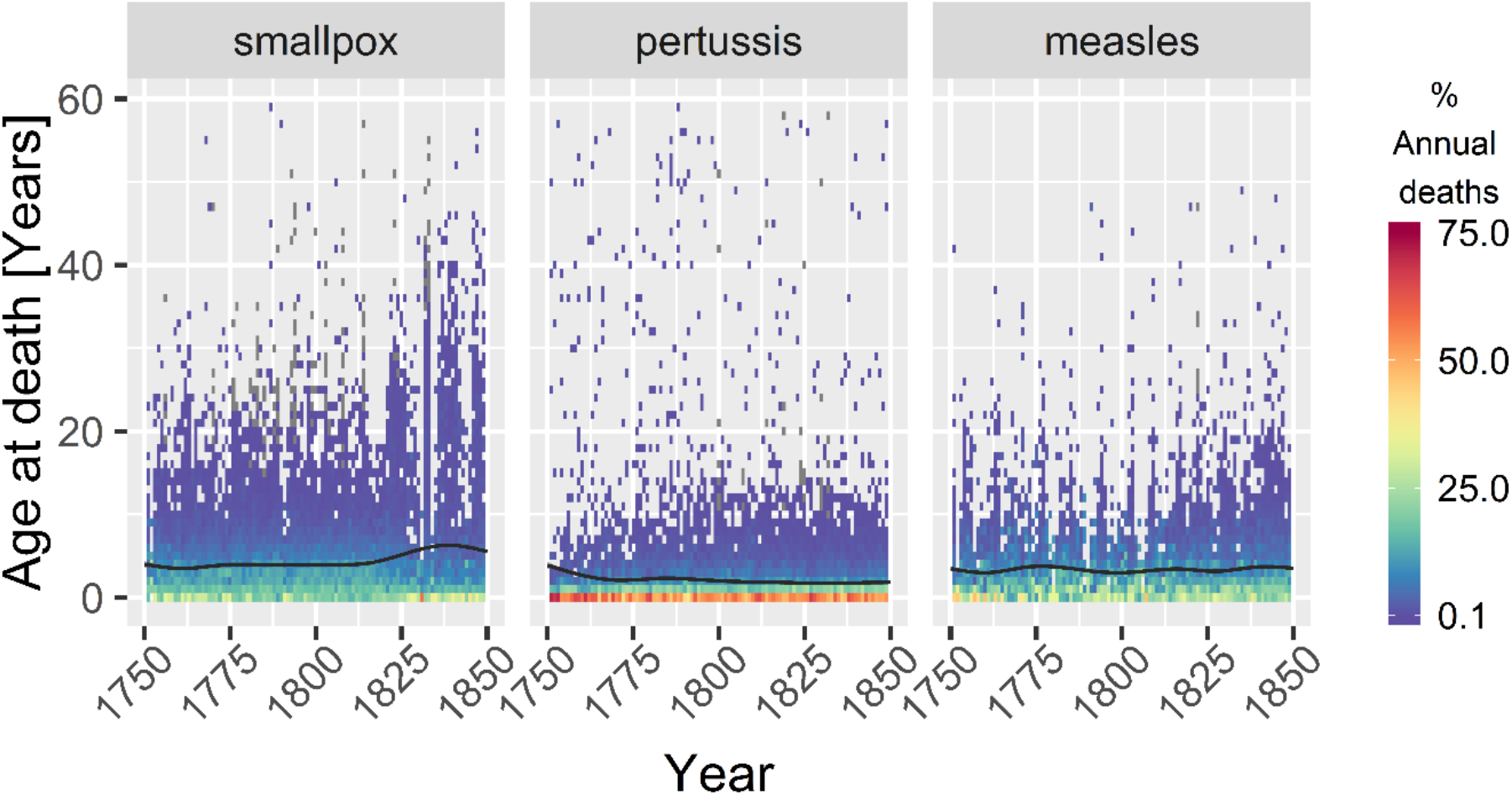
Distributions of ages at death for smallpox, pertussis and measles. Pertussis, affected the youngest children and smallpox the oldest, while measles was intermediate. Ages at death for individuals infected with smallpox showed a statistically significant increase from 1812 onwards, a decade after the start of vaccination, while the ages at death of pertussis declined over time and those of measles showed little overall change. Fitted lines are the result of best fitting GAM (Table S2; ±95CI). Data distributions and GAM derivatives are shown Fig. S5 A-D.

We tested the prediction that vaccination increased smallpox’ age at death and investigated whether this was different for the control infections pertussis and measles. We found strong statistical support for infection-specific changes in age at death over time (ΔAICc=-1641; Table S3B). Relative to the pre-vaccine era, smallpox’ age at death in the vaccine era *increased* by almost a year to 4.7 years (95CI: 4.7-4.8) and this increase was statistically significant (ΔAICc=-17.4; Table S3C; Fig. 3; Fig. S5 A-C). To estimate how quickly the introduction of vaccination in 1802 changed smallpox’ age at death, we fitted GAM and used the derivative to identify the year of increase, which for smallpox showed a statistically significant increase in age at death starting in 1812 (Fig. S5D).

In contrast, for pertussis, the mean age at death *decreased* in the vaccine era by half a year to 1.8 years (95CI: 1.8-1.8). This change was statistically significant and started from 1750 onwards (ΔAICc=-22.4; Table S3D; Fig. 3; Fig. S5 A-C). For measles the mean age at death increased by one month to 3.4 years, but this was not statistically significant (95CI: 3.4-3.5; ΔAICc=-1.3; Table S3E; Fig. 3; Fig. S5 A-C). Hence, the age at death from smallpox increased within a decade after introduction of the vaccine, while the infections without vaccination, pertussis and measles, showed either a decline or no change in age at death over time.

### 2.3 Periodicity of epidemics

We tested the prediction that vaccination increased the time period between epidemics and compared the change for smallpox with that for pertussis and measles. Wavelet analysis provided a good fit to the data (Fig. S6) and revealed that smallpox epidemics started off every 7 to 8 years and that outbreaks accelerated to every 4 to 5 years until 1812, at which point the periodicity in epidemics broke down and returned to approximately every 8 years (Fig. 2; Fig. 4A). In contrast, pertussis and measles started off with 7 to 8 years periods which gradually accelerated towards 4 years until the 1840’s (Fig. 4B & 4C). Thus, the epidemics of all childhood infections accelerated with time, but for smallpox, this acceleration broke down ten years after the introduction of vaccination.

**Fig. 4.**
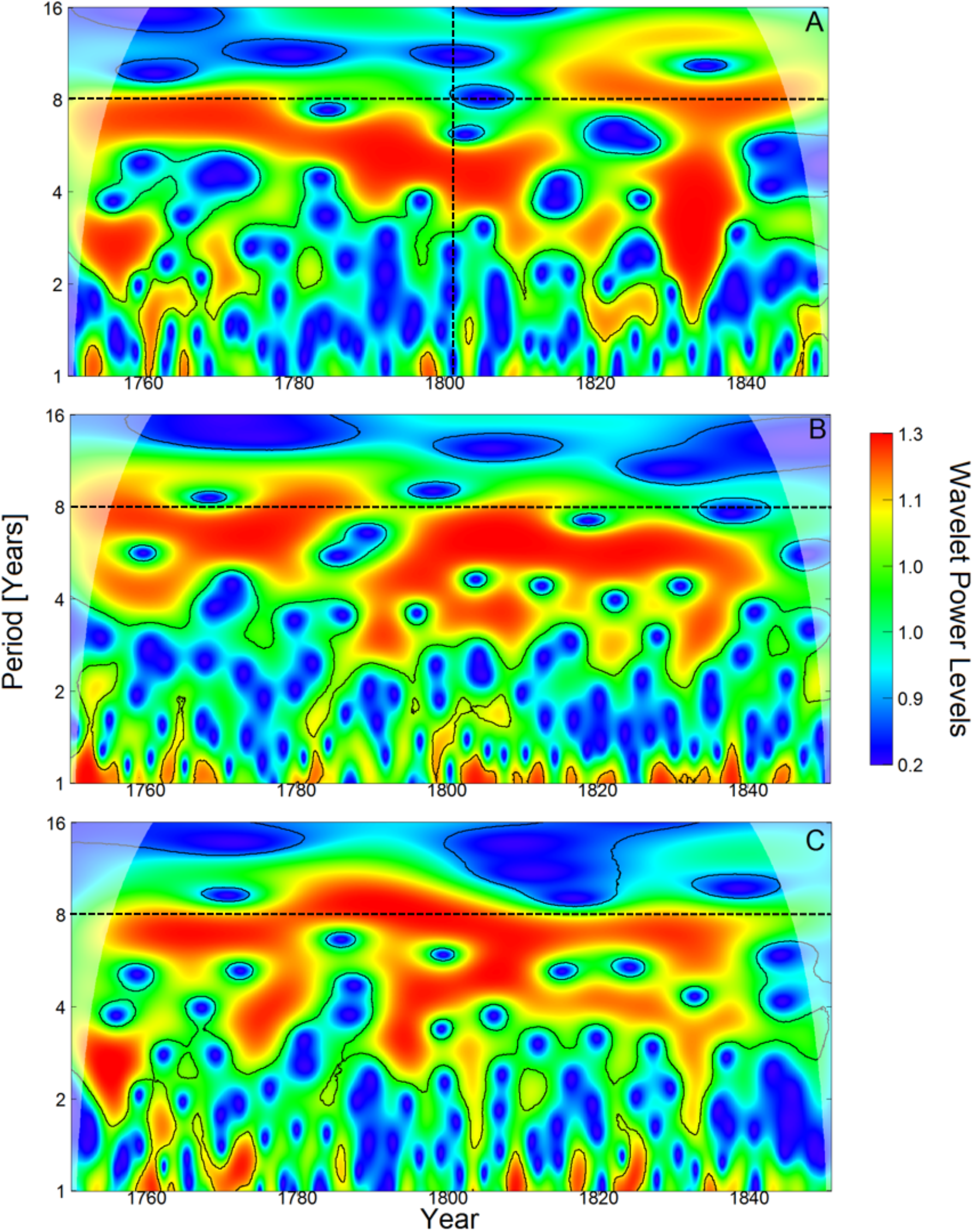
Wavelet power spectrum for (A) smallpox, (B) pertussis and (C) measles show that all infections start off with regular 7 to 8 year epidemics which accelerate with time. For smallpox the periodic structure breaks down in 1812 and returns to an 8 year periodicity, while for the infections without vaccination, pertussis and measles, the acceleration continues. Vertical dotted line denotes the introduction of the smallpox vaccine. Legends depict wavelet power levels with red referring to dominant periodicity and green or blue showing low levels of inferred periodicity. Solid black lines indicate significance at the 5% level. Shaded zones depict regions influenced by edge effects and should not be interpreted.

### 2.4 Fade-outs

To investigate whether vaccination affected the low-end of the infectious disease burden (in contrast to the high-end of the burden in the wavelet analyses), we analysed changes in fade-outs, i.e., the number of weeks per year without cause-specific deaths. We quantified the fade-outs at two spatial scales, nationwide and parish-specific, which gave consistent results (Tables S5 & S6 respectively). Changes in fade-out probabilities over time were infection-specific (−395.7<ΔAICc<-96.6; Fig. 5; Fig. S7 A & C; Tables S5A & S6A). For smallpox, fade-outs first decreased with time followed by an increase (−100.3<ΔAICc<-2.0; Fig. S7 A & C; Tables S5B & S6B). GAM derivatives revealed that nationwide fade-outs became rare after 1775 and increased significantly again from 1812 onwards (Fig. 5, Fig. S7B), while parish-specific fade-outs increased significantly in 1803, i.e., one year after the introduction of the smallpox vaccine (Fig. S7D). In contrast to smallpox, fade-outs for pertussis and measles decreased starting from 1750 and 1790 respectively (pertussis: -33.8<ΔAICc<-17.9; measles: -120<ΔAICc<-14.1; Table S5 C&D, Table S6 C&D). Thus, smallpox fade-outs increased within a decade after the introduction of vaccination, while those of the control infections pertussis and measles continued their decrease.

**Fig. 5.**
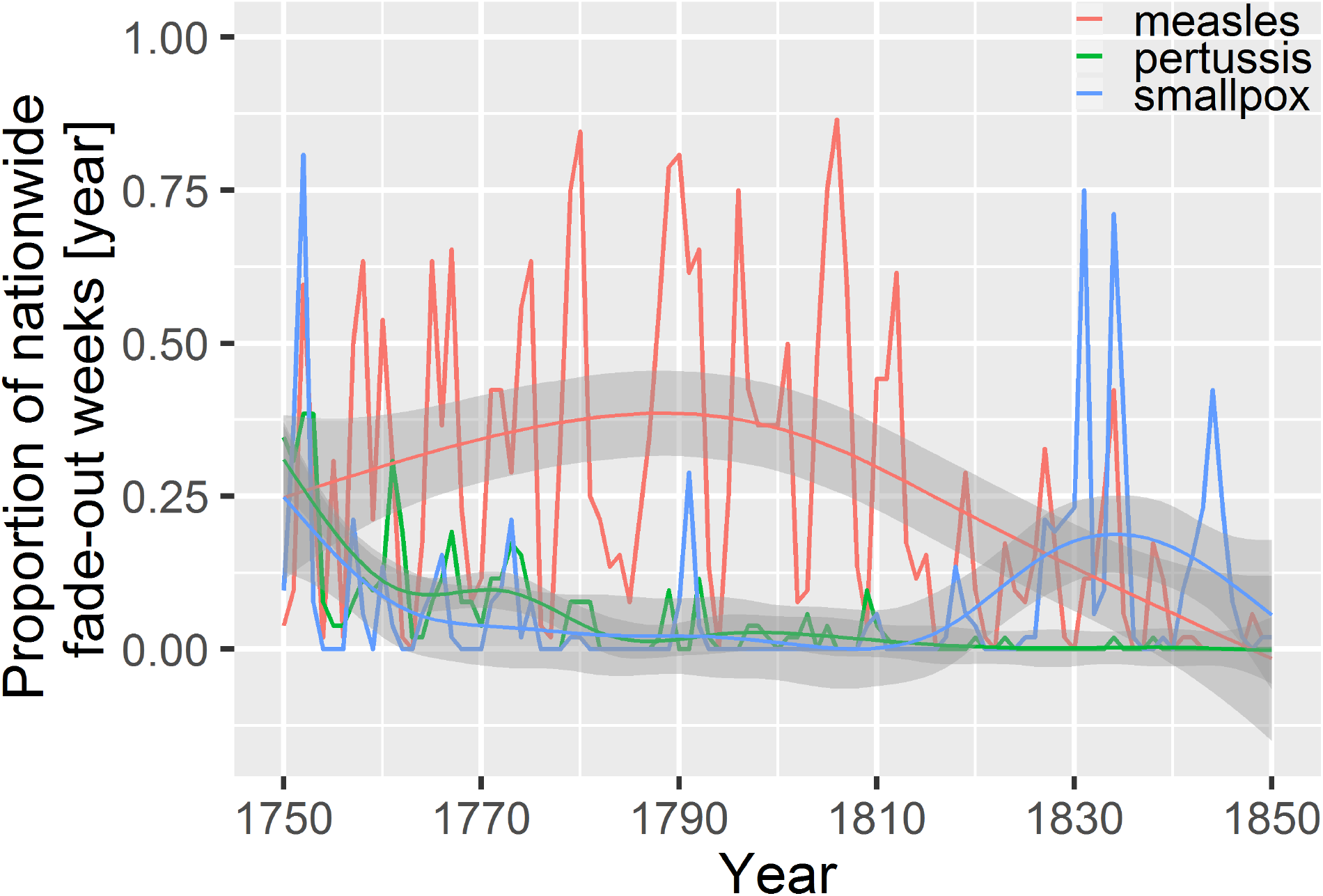
For smallpox, the annual proportion of weeks without infectious deaths decreased with time until 1812, after which it increased again. In contrast, for the infections without vaccination pertussis and measles, this probability decreased after 1802 with no sign of an increase. Lines show data and fitted lines represent GAM. For fade-outs at the parish level and for the GAM derivative see Fig. S7 A-D.

### 2.5 Reproduction numbers

Finally, we tested whether the transmission of childhood infections changed before vs. after the introduction of smallpox by comparing per era and per infection the reproduction numbers (R_0_), i.e., the number of new infections caused by a single infected individual in an entirely susceptible population (15). We did this using the mean age at infection (14) and time-series Susceptible-Infected-Recovered (TSIR) models (16), which gave consistent conclusions (Table 1; Fig. 6 A&B; Supplementary Information 5). Based on previous literature, we expected smallpox to have the lowest R_0_ of the three infections in our study (14), and we expected its R_0_ to decrease after the start of smallpox vaccination (15). Indeed, smallpox had the lowest R_0_ of the three infections, with a value of 6.1 in the pre-vaccine era (95CI: 6.1-6.3) which in the vaccine era decreased to 5.6 (95CI: 5.6-5.8; Fig. 6 A&B). Pertussis had the highest R_0_ of 10.4 in the pre-vaccine era (95CI: 9.9-10.8), which in the vaccine era increased to 15.0 (95CI: 14.9-15.9; Fig. 6 A&B). Measles had an intermediate R_0_ of 7.2 in the pre-vaccine era (95CI: 7.0-7.4), which increased in the vaccine era to 7.9 (95CI: 7.7-7.9; Fig. 6 A&B). Hence, smallpox had the lowest R_0_ which decreased after vaccination, while for the childhood infections without vaccination the R_0_ increased with time.

**Fig. 6.**
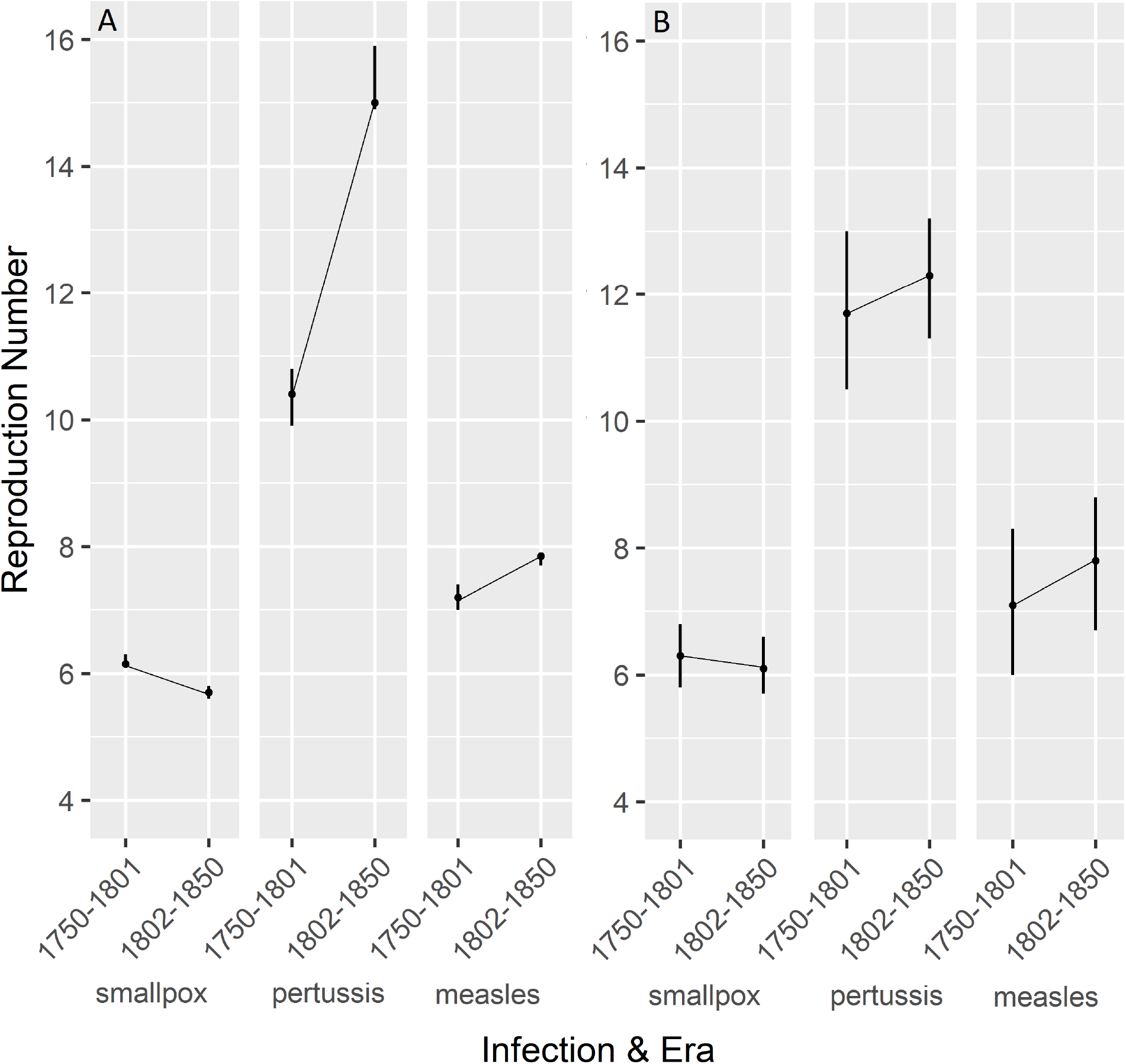
The reproduction number of smallpox decreased in the vaccine era, while for the control infections without vaccination, pertussis and measles, the reproduction number increased. (A) Reproduction numbers estimated using (A) mean age at infection and (B) TSIR models give consistent results but with larger 95CI (error bars).

## 3. Discussion

This study describes the epidemic dynamics of three childhood infections, smallpox, pertussis and measles, in a pre-vaccine 18^th^ and 19^th^ century European society. We show that these infections accounted for a large fraction of childhood mortality and that this mortality increased over time. However, within three years after the roll-out of the smallpox vaccine, smallpox mortality decreased abruptly, and within a decade, smallpox epidemiology showed all the theoretically predicted changes of the impact of vaccination with an increasing age at infection, periodicity, fade-outs and a decreasing transmission (R_0_). In contrast, most epidemiological characteristics of the childhood infections without vaccination, pertussis and measles, continued in the opposite direction to that of smallpox. Here we discuss three main implications of our study for our understanding of epidemic dynamics, public health interventions and the development of societies.

A first implication is that the epidemic of childhood infections in our population are slower relatively to those described in other pre-vaccine European societies (17–19). Little is known about pertussis and measles epidemiology during in the 18^th^ or 19^th^ century (for exceptions see (4, 20)), rendering comparisons difficult. However, for smallpox there is some information from other locations in 18^th^ century Europe. In 18^th^ or 19^th^ century London smallpox epidemics occurred at least twice as fast as in pre-vaccine Finland even though at that time London and Finland had similar population sizes (6, 21). However, Finland’s population density was far lower than that of London and with a patchy distribution, variables which decrease pathogen transmission, including in our population (15, 22). We believe this caused a relatively slow transmission, the slower epidemic dynamics and the many fade-outs. Indeed, in our study the estimates of R_0_ for all infections tended to be on the low side relative to the previously reported ranges in pre-vaccine 20^th^ century Europe: smallpox 5 - 6 vs. 3.5 - 6.6 (23), pertussis 10 - 15 vs. 10 – 17 (15) and especially measles for which the R_0_ of 7 to 8 is on the low side of the range of 6 to 27 (24). Hence, in historical Finland, epidemics and pathogen transmission were slower relative to those in pre-vaccine 20^th^ century European societies and this might largely be driven by Finland’s low population density and meta-population structure.

Second, the burden of childhood infections increased throughout the observation period. This is in contrast with other Nordic countries, where for example smallpox mortality decreased before the start of the vaccination programme (25). In the 18^th^ and 19^th^ century, Finland was characterized by minor improvements in public health. For example, in 1750, the country counted only one trained medical practitioner and by 1820 there were 373 hospital beds for 1.2 million inhabitants (26). At the same time however, Finland experienced rapid population growth (Fig. 1) and became increasingly connected to other European populations, which together with the limited public health can provide one explanation for Finland’s distinct increasing smallpox mortality.

Thirdly, our study shows an almost immediate response of smallpox epidemiology to the start of a vaccination programme, while other infections continued growing. After the introduction of vaccination, smallpox epidemics became irregular, but the interepidemic periods were at least twice as long as before the start of smallpox vaccination. During the same period, similar increases were reported in London (6, 21) and in Åland, an island nearby the Finnish mainland (27). Until now, it was not clear to what extent these changes were caused by vaccination or other socio-demographic improvements. In our study, the contrasting changes in epidemic dynamics between smallpox and the infections without vaccination, pertussis and measles, indicate that vaccination, rather than the development in standards of living and public health, is the main driver of these epidemiological changes. Our results here contrast with vaccination programmes against childhood infections in 20^th^ century Europe, which were also effective at reducing childhood mortality, but for which the vast majority of the decline occurred *before* the introduction of vaccines (5) and which emphasize the importance of the development in standards of living, hygiene and other public health improvements for the reduction of infection-associated childhood mortality.

One limitation of our study regarding the immediate impact of vaccination is that infections can interact. Hence the decrease in smallpox mortality could increase the number of susceptible individuals available for pertussis and measles, thereby creating contrasting dynamics between infections. While such interactions are theoretically expected (28), empirical evidence remains lacking. We believe that such interactions are possible, but for pertussis they cannot explain all of all the contrasting dynamics we found between smallpox because the increasing mortality and accelerating epidemics of pertussis started before the introduction of the smallpox vaccine (Fig. 3-5). Measles also showed accelerating epidemics before the introduction of the smallpox vaccine (e.g., Fig. 5), indicating other reasons for its increasing mortality, but the dynamics in Fig. 2 warrants further investigation. Interestingly, changes in smallpox epidemiology occurred abruptly, while the increase in pertussis and measles occurred gradually, indicating that interactions, if any, are not immediate but might be more complex, e.g., through immunity trade-offs (29) and these possibilities also warrant further study.

More generally, disentangling the contributions of vaccination from those of standards of living, hygiene and public health is key to informing public health interventions. Some claim that the decline in the burden of childhood infections in high-income countries can be attributed to general improvements in standards of living and health care rather than vaccination *per se* (30). Our results show that in Finland this claim is unlikely to be valid. In contrast to a background of increasing infectious disease mortality, Finland’s first vaccination programme was almost immediately effective at decreasing smallpox mortality, despite a growing and increasingly connected population. Highly transmissible childhood infections, such as measles, are on the rise in high income countries, an effect attributed in part to increasing vaccine hesitancy and providing information on the benefits of vaccination can decrease such hesitancy (30). Our results together with those of others (5) contribute to the robust evidence on the long-term effectiveness of vaccination programmes and hopefully provide incentive for the general population to make informed decisions on vaccinating children.

## 4. Materials and Methods

### 4.1 Study population and data

We used the data on births, deaths and causes of deaths gathered and digitized by the Finnish Genealogical Society available at http://hiski.genealogia.fi/historia/indexe.htm and last accessed on December 1^st^ 2017. The database contains 5,884,901 birth records and 3,490,737 death records collected between 1600 and 1948 in 507 parishes. For the purpose of this study, we focused on the years with the most complete birth and causes of death data, i.e., between 1750 and 1850. We selected those parishes without missing years in the data and below the Arctic circle (66°33′47.1’’). The population above the Arctic Circle is different in many ways: they were scarcely inhabited by Saami, a nomadic population of reindeer herders who, in contrast to the more agricultural south, depended for their livelihood on herding, fishing and hunting (31). Based on these criteria, we used data from 213 parishes (Fig. 1A).

In the historical records, the causes of death were identified or recorded by parish priests. The records contained 51,075 causes of death, written in Swedish, Finnish or German. We identified the causes of interest first by classifying causes according to their spelling similarity and then by their synonyms in different languages following (32). This was done by two investigators independently (M.B. and T.K.) and resulted in a consistent outcome. A death cause was missing for 104,925 records (9%) and 693 records (0.06%) missed an age at death. These were excluded, which together with the above selection criteria, resulted in a database with 1,693,056 birth records and 1,190,627 death records.

We chose these three infections because they are readily infections: smallpox and measles have characteristic rashes and pertussis has a distinct cough. The three pathogens also share similar characteristics: they are transmitted either through direct contact or via droplets and have relatively similar generations times of two weeks for smallpox and measles and four weeks for pertussis (14).

### 4.2 Analyses

We performed all analyses using R version 3.5.1 (33)and few performed all analyses with both generalized linear models (GLM), general additive models (GAM). These models have in common that to identify the statistical significance of predictor variables, we used model selection (34, 35) based on the second order Akaike Information Criterion (AICc). In brief, this is a hypothesis-based approach that generates, given a global model, subset models (varying in predictors) that best fit the data. Better fitting models are indicated by their lower AICc, which is a continuous variable with models within 4 ΔAICc being plausible and becoming increasingly equivocal up to 14 ΔAICc, after which they become implausible (35).

#### 4.2.1 Time series of infectious disease mortality

In the analyses of infectious disease mortality, the aim is to identify if and when smallpox vaccination affected smallpox mortality. To this end, we followed the same approach as performed previously (22), running three binomial GLMs for smallpox, pertussis and measles. We did this with both the functions ‘glmer’ of the package ‘lme4’ (36) and with ‘glmmTMB’ of the package ‘glmmTMB’ (37), which always gave consistent results. The dependent variable was whether or not an individual had died from the infection of interest. Annual population size was used as an offset, i.e., all individuals in the population who had not died from the infection of interest received 0 (22). We used a binomial error distribution, we accounted for spatial clustering by including parish identity as a random intercept and corrected for overdispersion by including an observation level random effect.

We analysed the time series with death year as the predictor variable. The data show many non-linear dynamics (Fig. 2) and here we wanted to identify the year of major change in infection mortality. To this end, we used two ways: (i) gradually, with a GLMs including year up to a quadratic term or (ii) abruptly changes using threshold GLM (38) and (iii) using GAM and their derivative to identify their inflection point (see supplementary information 2 for more detailed explanation). In brief, threshold GLMs estimate a year in which there is a sudden change in the dependent variable by testing, using the AICc, the fit on the data of a large number of GLMs, each one with a different threshold year. We computed the ‘confidence’ around the threshold year by inverting the 4 ΔAICc acceptance region of the model fit (38).

#### 4.2.2 Age at infection

To test the prediction that vaccination increases the age at infection, we first estimated mean ages at death in pre-vaccination and vaccination eras using arithmetic and gamma distributed means, which showed consistent results. We then investigated changes in ages at death across time using GLM and GAM with a gamma error distribution. To identify the time point of increase in age at infection we used the GAMs’ first derivative and threshold GLM, which gave consistent results. In all models, we accounted for the spatial clustering of the data by including parish identity as random intercept (ΔAICc=-4027; Table S1E) and for temporal autocorrelation of the data by including death year as an auto-regressive factor of order 1 (ΔAICc=-1039; Table S1E). We explain these methods in more detail in Supplementary Information 2.

### 4.2.3 Periodicity of epidemics

To detect whether the epidemics showed cyclic dynamics, we conducted wavelet analyses. In brief, wavelet analysis decomposes time series data using functions (wavelets) simultaneously as a function of both period and time (39). We pooled the data into monthly intervals and tested periods between one and sixteen years. To determine statistical significance, we compared the periodicity of the data with that of 1000 ‘white noise’ datasets with significance at the 5% level. We give more detailed information and show the wavelet fits on the data in Supplementary Information 3.

### 4.2.4 Fade-outs

We quantified fade-outs as number of weeks per year without infectious deaths and analysed changes over time using GLM with a quasibinomial error distribution. We also performed the analyses using fade-outs per two weeks and per month, which led similar conclusions (results not shown). Other analyses specifications were as in section 4.2.1 and we give more explanation in Supplementary Information 4.

### 4.2.5 Reproduction numbers

To test whether vaccination decreased the transmission of smallpox, we estimated the reproduction numbers. We used two complementary approaches to estimate R_0_, one using age at infection and one using TSIR models. TSIR models fitted the data well (Fig. S8) and both approaches gave consistent results (Fig. 6 A &B). We explain the methods in detail in Supplementary Information 5.

## Data Availability

All data produced are available online at: https://hiski.genealogia.fi/historia/indexe.htm

## Acknowledgements

We are grateful to the many volunteers of Genealogical Society of Finland who collected the dataset from church records and to useful discussions and feedback from the Nordemics Consortium, Alex Becker, Vérane Berger, Terhi Honkola, Rob Lynch, Kimmo Pokkinen and Angi Rösch. We thank the funding from the Academy of Finland (TK: 278751; VL: 292368), NordForsk (104910), the Finnish Cultural Foundation (TK) and the Ella & Georg Ehrnrooth Foundation (MB).

