## Supplementary material for "The epidemic dynamics of three childhood infections and the impact of first vaccination in 18^th^ and 19^th^ century Finland": Table S1-S6; Fig S1-S8

### Supplementary information to:
### The epidemic dynamics of three childhood infections and the impact of first vaccination in 18th and 19th century Finland

Michael Briga[1,2*], Tarmo Ketola[3], Virpi Lummaa[1]

[1] Department of Biology, University of Turku, Turku, Finland

[2] Infectious Disease Epidemiology Group, Max Planck Institute for Infection Biology, Berlin, Germany.

[3] Department of Biological and Environmental Science, University of Jyväskylä, Jyväskylä, Finland

#### Table of contents

1. **Data validation and childhood mortality rates** **2**
   Fig. S1- S4, Table S1
2. **Time series of infectious disease mortality** **6**
   Table S2
3. **Ages at death analyses** **7**
   Table S3, Fig. S5
4. **Wavelet analyses** **11**
   Fig. S6
5. **Fade-out analyses** **12**
   Table S5 & S6, Fig. S7
6. **Reproduction numbers** **16**
   Fig. S8
7. **References** **18**

**Supplementary Information 1: Data validation**

In the historical records, the causes of death were recorded by parish priests. Misdiagnosis is possible, but to limit this source of error, we used three readily identifiable infections. Chickenpox could be distinguished from smallpox as early as in the 17th Century (Fenner et al. 1988) and chickenpox case fatality rates were low (Hussey et al. 2017) and hence confusing the cause of death between smallpox mortality and chickenpox is unlikely. The records contained 51,075 causes of death, written in Swedish, Finnish or German. We identified the causes of interest first by classifying causes according to their spelling similarity and then by their synonyms in different languages following (Vuorinen 1999). This was done by two investigators independently (M.B. and T.K.) and resulted in a consistent outcome. A death cause was missing for 104,925 records (9%) and 693 records (0.06%) missed an age at death. These were excluded, which together with the selection criteria mentioned in the methods section of the manuscript, resulted in a database with 1,693,056 birth records and 1,190,627 death records.

Since we use subset of the data, we validated the database as follows. First, we estimated the % of birth records for which there was a death record, which shows that for those cohorts with up to 80 years of monitoring, almost 100% of the birth records have a death record (Fig. S1). Second, the correlation between the time series of births and deaths between the database and those of Finland as a whole from the Official Statistics of Finland was high at 0.97 and 0.94 respectively (Fig. S2; Official Statistics of Finland 2018). Third, per capita birth and mortality rates were high at 0.040 (interannual SD: 0.004) and 0.030 (interannual SD: 0.007) respectively, which values were consistent with those from other demographic databases in the same period (Kannisto et al. 1999; Scranton et al. 2016). Fourth, age-specific mortality rates (Fig. S3) are close to age-specific survival or mortality curves from other demographic studies (Turpeinen 1979; Bolund et al. 2015; Scranton et al. 2016). Fifth, the population size in this database increased from 212,850 to 705,701 individuals. Finland's population at that time increased from roughly 400,000 to 1,600,000 inhabitants (Official Statistics of Finland 2018). The data thus capture 50% of Finland's population in this period (interannual SD: 5%).

*Fig. S1 Estimation of the birth records for which there was a death record, showing that for the birth cohorts with 80 years of monitoring, close to 100% of birth records were covered. Noteworthy, in the last year of observation, 1850, 25% of the births also had a death record, which matches the mortality rates reported from other datasets for that period (Turpeinen 1979; Kannisto et al. 1999; Scranton et al. 2016).*

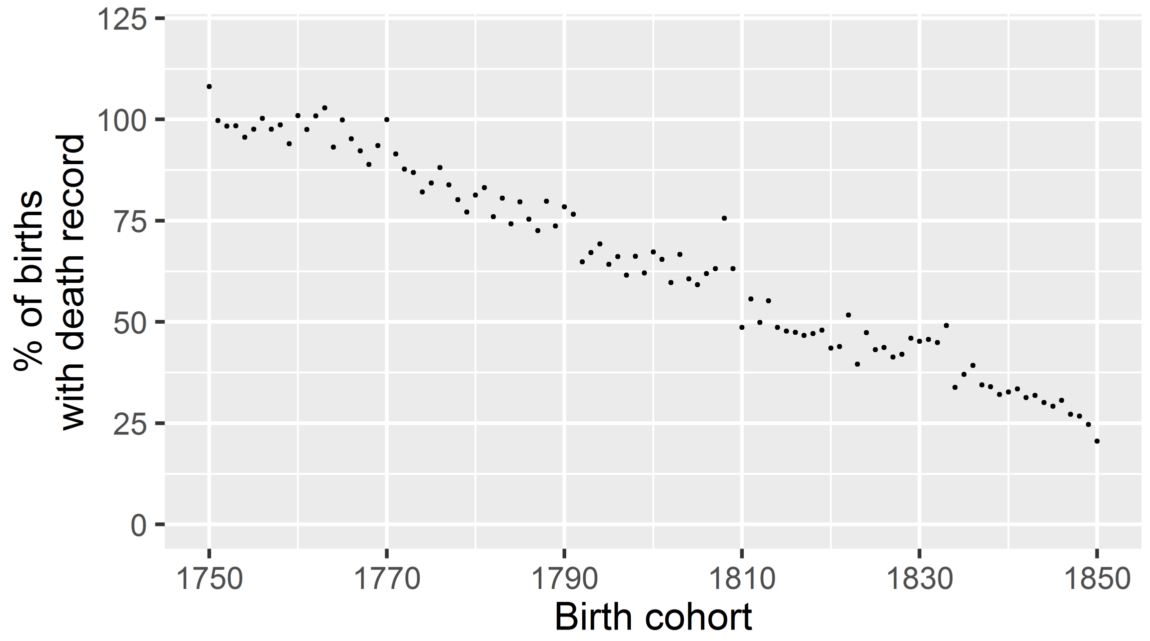


*Fig. S2 Time series of (A) births and (B) death of our dataset are highly correlated with the nationwide data at 0.97 and 0.94 respectively.*

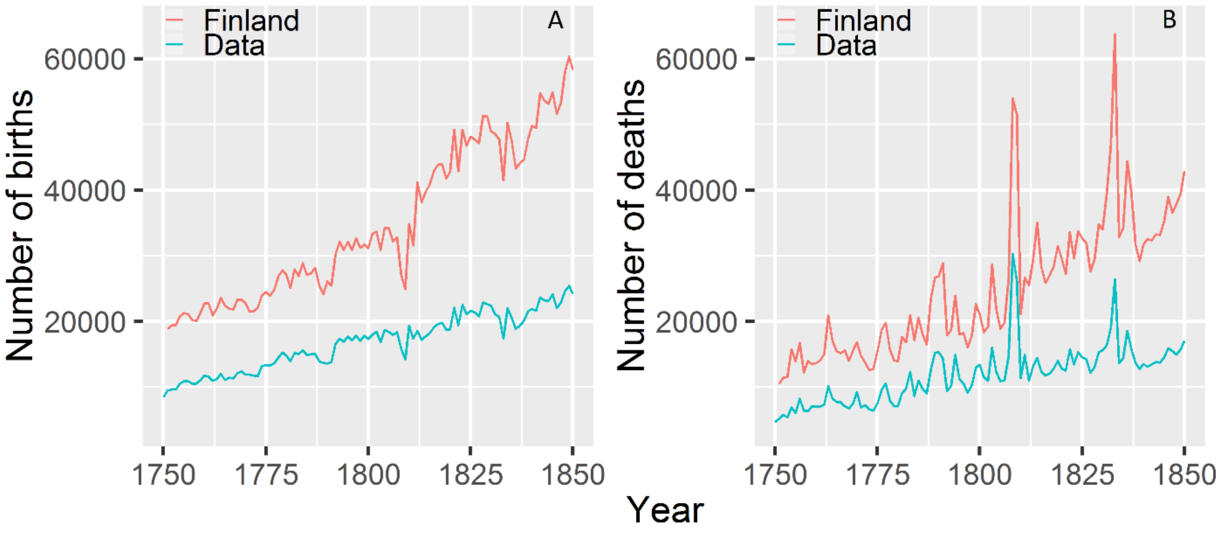

*Fig. S3 Dynamics of childhood mortality per birth cohort. Different lines represent years of monitoring since the birth of a cohort. Lines with longer years of monitoring stop earlier because the data monitoring stopped in 1850.*

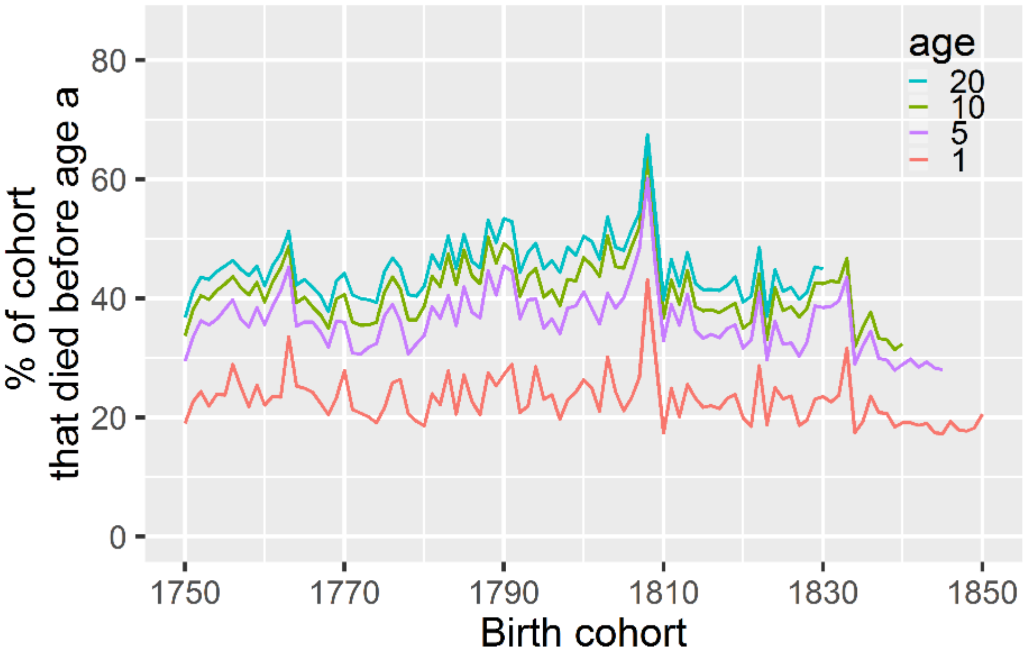


*Fig. S4 Proportion of registered deaths that could be attributed to the three childhood infections smallpox, pertussis and measles. Different lines represent years of monitoring since the birth of a cohort, with monitoring stopping in 1850.*

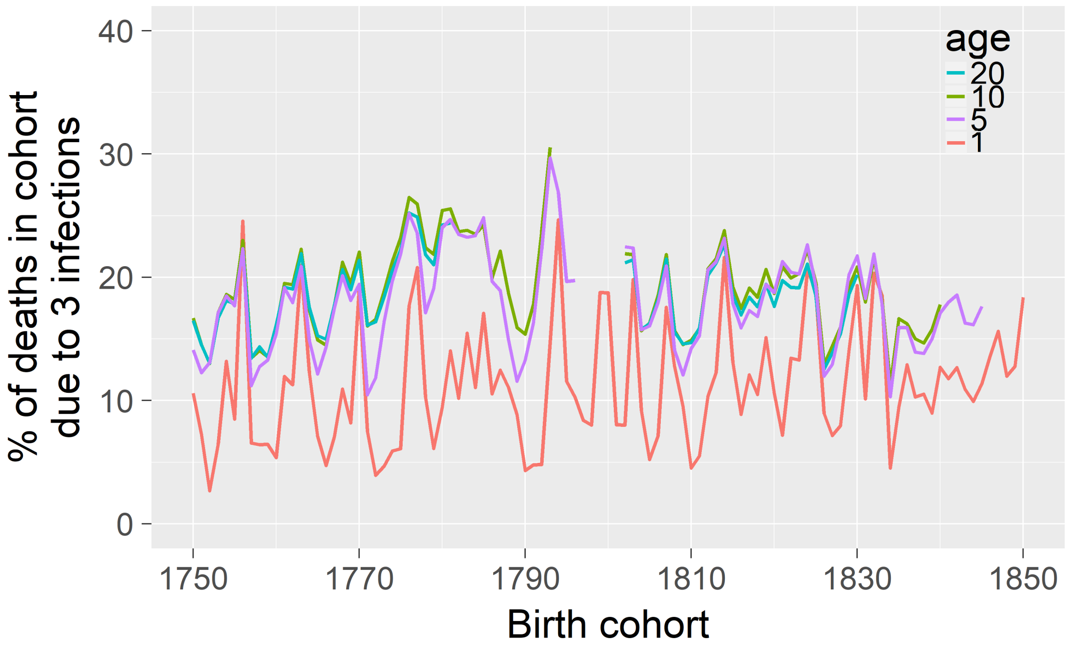

*Table S1. The mean childhood mortality rates (%, ± inter birthcohort sd) per birth cohort of all registered births in our database. Pre-vaccine and vaccine eras refer to before and after the introduction of the smallpox vaccine in 1802. Childhood mortality was high and over 40% by age 10 years in both eras, with an improvement between eras ranging from 0.7% to 1.5%. The contributions of the three childhood infections to childhood mortality remained high and close to 20% by age 10, with a decline of 1.6% for children before versus after the start of smallpox vaccination, i.e. the pre-vaccine and the vaccine eras respectively. Note that for children until age 1, the change is in the opposite direction: there is an 1.5% increase in mortality due to the three infections after the introduction of the smallpox vaccine. This is due to the changes in the prevalence of childhood infections: while smallpox mortality decreased, pertussis mortality increased (Fig. 2) and pertussis age at death is younger than smallpox age at death (Fig. 3), causing an opposite trend in mortality differences at age 1 relative to age 5 or age 10.*

| | **Childhood mortality** | | | **Contribution of the 3 infections to childhood mortality** | | |
|---|---|---|---|---|---|---|
| **era** | **pre-vaccine** | **vaccine** | **mortality** | **pre-vaccine** | **vaccine** | **mortality** |
| **age [years]** | **1750-1801** | **1802-1850** | **difference** | **1750-1801** | **1802-1850** | **difference** |
| 1 | 23.7 (3.1) | 22.2 (4.7) | -1.5 | 10.6 (5.4) | 12.1 (4.4) | 1.5 |
| 5 | 36.8 (3.8) | 35.5 (6.4) | -1.3 | 18.4 (4.6) | 17.6 (3.2) | -0.8 |
| 10 | 41.4 (4.1) | 40.7 (6.6) | -0.7 | 19.8 (4.2) | 18.2 (2.9) | -1.6 |

**Supplementary Information 2: Time series of infectious disease mortality**

*Table S2. Dynamics of infectious disease mortality from (A) smallpox, (B) pertussis and (C) measles. Models are GLMs fitted on individual level data using a binomial error distribution and correct for temporal and spatial autocorrelation.*

| Model | Intercept | year | $year^2$ | df | AICc | delta AICc | weight |
|---|---|---|---|---|---|---|---|
| **(A) smallpox** | | | | | | | |
| threshold | -2.09 | 0.55 | -1.40 | 6 | 182154.0 | 0.0 | 0.56 |
| quadratic | -3.18 | -0.76 | -0.36 | 6 | 182155.0 | 1.0 | 0.37 |
| linear decrease | -3.55 | -0.40 | | 5 | 182159.7 | 5.7 | 0.03 |
| no change | -3.36 | | | 4 | 182163.0 | 9.0 | 0.03 |
| **(B) pertussis** | | | | | | | |
| linear increase | -3.67 | 0.24 | | 5 | 120321.7 | 0.0 | 0.71 |
| accelerating increase | -3.66 | 0.22 | -0.02 | 6 | 120323.6 | 1.9 | 0.27 |
| no change | -3.78 | | | 4 | 120328.3 | 6.6 | 0.03 |
| **(C) measles** | | | | | | | |
| quadratic upward opening | -5.07 | 0.59 | 0.47 | 6 | 79959.7 | 0.0 | 0.95 |
| no change | -4.64 | | | 4 | 79966.5 | 6.8 | 0.03 |
| linear increase | -4.58 | 0.12 | | 5 | 79968.1 | 8.4 | 0.01 |

#### Supplementary Information 3: Ages at death analyses

We tested the prediction that vaccination increased the age at infection in three ways (see below). For all these analyses we used age at death, quantified up to annual resolution. The first administration of a smallpox vaccine in Finland was reported in 1802, and we included all death dates occurring before January 1st 1802 as pre-vaccine deaths and hence we determined in the pre-vaccine vs. vaccine eras as 1750-1801 and 1802-1850. Below here we explain the three approaches, which we all performed in R (R Core Team 2019) following Zuur et al. (2009) and Woods (2017).

First, we estimated mean ages at death in the pre-vaccination vs. vaccination eras. We did this using three different approaches: (i) arithmetic means, (ii) fitting gamma distributions and (iii) using generalized linear models (GLM) with Gamma error distribution. We obtained arithmetic means with the function 'mean' of the package 'base' (R Core Team 2019) and estimated the confidence intervals around the arithmetic means by bootstrapping with the function 'boot' from the package 'boot' (Canty and Ripley 2017). We fitted Gamma distributions with the function 'fitdist' of the package 'fitdistrplus' (Delignette-Muller and Dutang 2015). GLM with a gamma error distribution were fitted the function 'glmer' of the package 'lme4' (Bates et al. 2015). These three approaches gave consistent results (results not shown).

We then investigated changes in ages at death between the pre-vaccine and vaccine eras using generalized linear models (GLM) and generalized additive models (GAM) with a Gamma error distribution and a log link function. GAM are better to quantify possible complex non-linear dynamics but GLM allow a more straightforward interpretation of simple non-linear dynamics, for example by testing interactions of infections-specific changes with time. Both approaches gave consistent results (Table S3 vs. Fig. S5C: for smallpox and pertussis the shapes are consistent; for measles there is no statistically significant change with time). We fitted GLM with the function 'glmmTMB' of the package 'glmmTMB' (Brooks et al. 2017), which gave results consistent with those fitted with 'glmer' of the package 'lme4' (Bates et al. 2015) and with the Bayesian models fitted with 'brm' of the package 'brms' (Bürkner 2017; results not shown). We compared model fits based on the second order Akaike Information Criterion (Burnham and Anderson 2002; Burnham et al. 2011) as implemented with the function 'AICc' of the package 'MuMIn' (Barton 2019). We controlled model residuals with the function 'simulateResiduals' of the package 'DHARMa' (Hartig 2019) and these fulfilled all assumptions homoscedasticity without overdispersion, zero-inflation, outliers, temporal or autocorrelation.

In all glmmTMB models, we corrected for temporal autocorrelation by including year as an auto-regressive factor of order 1 (AR1; ΔAICc=-1039 best fitting model in Table S3B relative to the same model without temporal autocorrelation). We accounted for the spatial clustering of the data by including parish identity as random intercept. This improved the model fit (ΔAICc=-4027, best fitting model in Table S3B relative to the same model without spatial clustering). Model residuals did not retain any statistically significant temporal or spatial autocorrelation as checked with 'testTemporalAutocorrelation' of the package 'DHARMa' (Hartig 2019), with 'acf' of the package 'stats' (R Core Team 2019) and with 'testSpatialAutocorrelation' using Moran's I of the package 'ape' (Paradis and Schliep 2019) on (un)conditional residuals.

To identify in which year there was an increase in age at death accounting for the non-linear dynamics in age as death with time, we fitted GAM with the function 'gamm' of the package 'mgcv' (Woods et al. 2016; Woods 2017). In these models, age at death was the dependent variable and death year the predictor variable, smoothened with cubic splines 'cr', as they fitted the data best ($-589<\Delta AICc<-5.5$), but other smoothers resulted in the same conclusions (results not shown). We used a gamma error distribution, included parish identity as random intercept and included death year as an AR1 temporal autocorrelation structure. We then identified the year of increase in age at death using the GAM's inflection point based on the GAM derivative, i.e., the age at death increases on the year that the derivative crosses zero to become positive (Fig. S5D). We calculated the GAM derivative using the function 'fderiv' of the package 'gratia' (Simpson 2019). This increase becomes statistically significant when the confidence around the derivative does not overlap with zero. We estimated the confidence following (Ruppert et al. 2003), which in brief consists of a 10,000 posterior simulations on 200 points along the fitted GAM and taking the lower 2.5% and upper 97.5% of that distribution at each of these points.

*Table S3 Strong model support for differences in ages death between infections in (A) the pre-vaccine and (B) over the study period showing (C) an increase for smallpox, (D) a decrease for pertussis, but (E) no statistically significant change for measles. Models are GLMs with a gamma error distribution, parish identity as random intercept and correct for temporal autocorrelation by including year of death as an auto-regressive factor of order 1. inf = infection; + = >1 coefficient and hence not shown in table S3B, but shown in table S3C, D and E.*

| **Terms** | | | | | | **Model Fit** | | | | **Shape** |
|---|---|---|---|---|---|---|---|---|---|---|
| **(A) Three infections 1750-1801 (pre-vaccine era)** | | | | | | | | | | |
| **intercept** | **NA** | **NA** | **inf** | **NA** | **NA** | **df** | **AICc** | **dAICc** | **weight** | |
| 1.16 | | | + | | | 7 | 265108 | 0 | 1.0 | NA |
| 1.18 | | | | | | 5 | 267605 | 2497 | 0.0 | NA |
| **(B) Three infections 1750-1850** | | | | | | | | | | |
| **intercept** | **year** | **year2** | **inf** | **year*inf** | **year2*inf** | **df** | **AICc** | **dAICc** | **weight** | **shape** |
| 1.14 | 0.02 | 0.04 | + | + | + | 13 | 586177 | 0 | 1.0 | NA |
| 1.11 | 0.03 | 0.06 | + | + | | 11 | 586201 | 23.9 | 0.0 | NA |
| 1.18 | 0.00 | | + | + | | 10 | 586218 | 40.5 | 0.0 | NA |
| 1.13 | 0.01 | 0.05 | + | | | 9 | 587818 | 1640.9 | 0.0 | NA |
| 1.19 | | | + | | | 7 | 587826 | 1648.5 | 0.0 | NA |
| 1.19 | -0.01 | | + | | | 8 | 587827 | 1650.3 | 0.0 | NA |
| 1.17 | -0.05 | | | | | 6 | 600664 | 14486.9 | 0.0 | NA |
| 1.18 | | | | | | 5 | 600668 | 14490.5 | 0.0 | NA |
| **(C) Smallpox** | | | | | | | | | | |
| **intercept** | **year** | **year2** | **NA** | **NA** | **NA** | **df** | **AICc** | **dAICc** | **weight** | **shape** |
| 1.39 | 0.16 | 0.07 | | | | 7 | 336724 | 0 | 1.0 | upward opening |
| 1.47 | 0.13 | | | | | 6 | 336733 | 9.1 | 0.0 | increase |
| 1.46 | | | | | | 5 | 336741 | 17.4 | 0.0 | no change |
| **(D) Pertussis** | | | | | | | | | | |
| **intercept** | **year** | **year2** | **NA** | **NA** | **NA** | **df** | **AICc** | **dAICc** | **weight** | **shape** |
| 0.57 | -0.14 | 0.07 | | | | 7 | 148002 | 0 | 1.0 | accelerating decline |
| 0.65 | -0.17 | | | | | 6 | 148009 | 6.7 | 0.0 | decline |
| 0.70 | | | | | | 5 | 148025 | 22.4 | 0.0 | no change |
| **(E) Measles** | | | | | | | | | | |
| **intercept** | **year** | **year2** | **NA** | **NA** | **NA** | **df** | **AICc** | **dAICc** | **weight** | **shape** |
| 1.17 | 0.03 | | | | | 6 | 99068 | 0 | 0.5 | increase |
| 1.17 | | | | | | 5 | 99069 | 1.3 | 0.3 | no change |
| 1.17 | 0.03 | 0.00 | | | | 7 | 99070 | 1.9 | 0.2 | upward opening |

*Fig. S5 In the vaccine era (1802-1850; vac; dotted lines) smallpox' ages at death (blue) increased relative to the pre-vaccine era (1750-1801; prevac; solid lines), while this was not the case for the control infections pertussis and measles (green and red). (A) Distributions and (B) cumulative distributions (B) of ages at death per infection. (C) Best fitting GAM models and (D) the GAM derivative for smallpox showing the significantly increasing age starting in 1812. Shaded areas show 95CI.*

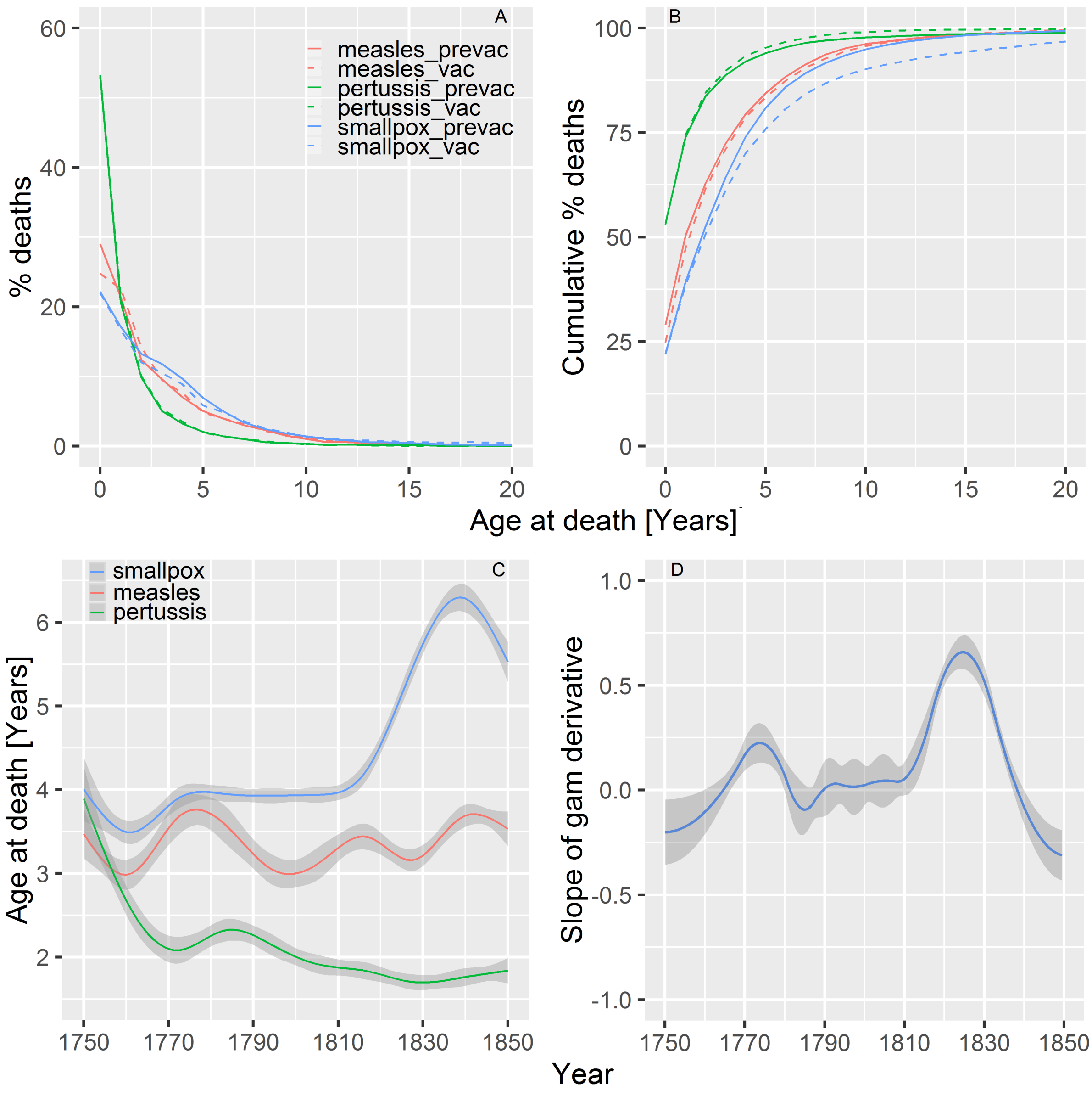

#### Supplementary Information 3: Wavelet analyses

We performed wavelet analyses following (Cazelles et al. 2007; Roesch and Schmidbauer 2018). In brief, we fitted a Morlet wavelet with the function ‘analyze.wavelet’ using the package ‘WaveletComp’(Roesch and Schmidbauer 2018). We pooled the data into monthly intervals and log-tranformed (+1) the time series. Analyses without or with a range of normalizing the data (loess.span’) gave consistent results. We controlled the fit of the wavelets by reconstructing the time series in Fig. S6.

*Fig. S6 Match between the reconstructed epidemics (red) based on the periodicities of the wavelet analyses and the epidemic dynamics of the data (black) for (A) smallpox, (B) pertussis and (C) measles.*

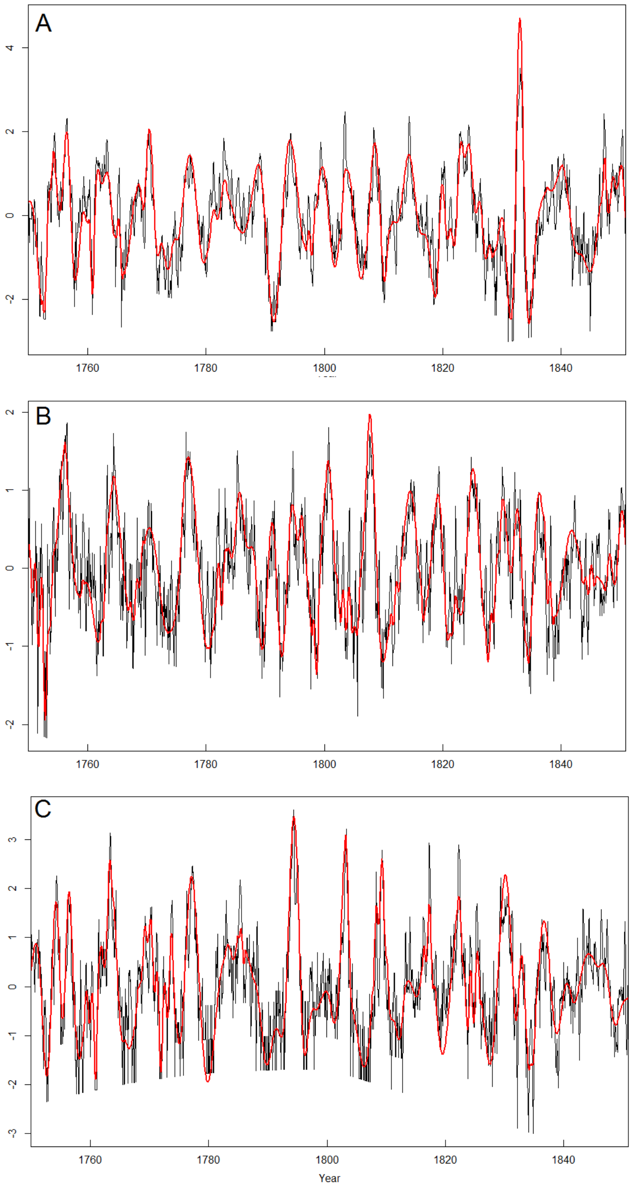

**Supplementary Information 4: Fade-outs analyses**

Consistent with the analyses for age at death, we studied the temporal dynamics in the number of fade-out weeks per year we using both GLM and GAM and both approaches gave consistent conclusions (Table S5 vs. Fig. S7A; Table S6 vs. Fig S7C). We fitted GLM with a betabinomial error distribution in the function 'glmmTMB' from the package 'glmmTMB' (Brooks et al. 2017). In these models, the dependent variable is the number of weeks per year with or without infectious deaths (binomially coded) and year is the predictor variable. We quantified the number of fade-out weeks per year at two spatial levels, national and parish-specific. The nationwide quantification is often used in the literature (Ferrari et al. 2013), but the regional analyses can account for spatial clustering of the fade-outs (see below). Note that the difference in fade-out values between both levels reflect spatio-temporal asynchrony between epidemics in different parishes (Rohani et al. 1999; Grenfell et al. 2001). For the nationwide fade-out analyses, we did not include parish identity as a random term as that unavoidably generates a dependent variable at the parish level. For the parish-specific fade-out analyses we included parish identity as a random intercept. There was a statistically significant temporal autocorrelation over a two-year time lag and we accounted for this by including year as an auto-regressive factor of order 1 (AR1), which improved the model fit (nationwide: AICc=-34.1 in the best fitting model in Table S5A; parish-specific: AICc=-1280 in the best fitting model in Table S6A). To identify more precisely the year of change in fade-out weeks we fitted GAM with a quasibinomial error structure and cyclic cubic splines 'cc' (as they fitted the data best, but other smoothers gave consistent results (results not shown) and then estimated the GAM derivative as explained in supplementary information 3. We performed model residuals checks and estimations of confidence intervals of model parameters as explained in in supplementary information 3 and model residuals fulfilled all assumptions and were without temporal or spatial autocorrelation.

*Table S5 Nationwide fade-out analyses support (A) infections-specific changes in fade-out probabilities over time with (B) for smallpox a parabolic quadratic association while (C) and (D) pertussis and measles show a declining probability. Models are GLM with a betabinomial error structure and correct for temporal autocorrelation. Other abbreviations as in Table S1.*

| Terms | | | | | | Model Fit | | | | Shape |
|---|---|---|---|---|---|---|---|---|---|---|
| **(A) Three infections** | | | | | | | | | | |
| **intercept** | **year** | **year2** | **inf** | **year*inf** | **year2*inf** | **df** | **AICc** | **dAICc** | **weight** | |
| -0.58 | -0.85 | -0.91 | + | + | + | 12 | 1407.9 | 0 | 1.0 | NA |
| -1.10 | -0.54 | -0.14 | + | + | | 10 | 1470.0 | 62.1 | 0.0 | NA |
| -1.19 | -0.48 | | + | | | 7 | 1504.6 | 96.6 | 0.0 | NA |
| -1.15 | -0.49 | -0.04 | + | | | 8 | 1506.5 | 98.6 | 0.0 | NA |
| -1.19 | | | + | | | 6 | 1521.9 | 114.0 | 0.0 | NA |
| -1.47 | -0.68 | | + | + | | 8 | 2315.7 | 907.8 | 0.0 | NA |
| **(B) Smallpox** | | | | | | | | | | |
| **intercept** | **year** | **year2** | **NA** | **NA** | **NA** | **df** | **AICc** | **dAICc** | **weight** | **shape** |
| -5.99 | 0.16 | 1.41 | | | | 6 | 400.3 | 0 | 0.6 | upward opening |
| -4.49 | 0.10 | | | | | 4 | 402.2 | 2.0 | 0.2 | increase |
| -4.49 | | | | | | 4 | 402.2 | 2.0 | 0.2 | no change |
| **(C) Pertussis** | | | | | | | | | | |
| **intercept** | **year** | **year2** | **NA** | **NA** | **NA** | **df** | **AICc** | **dAICc** | **weight** | **shape** |
| -4.47 | -1.76 | | | | | 4 | 305.1 | 0 | 0.8 | decline |
| -4.34 | -1.57 | 0.13 | | | | 6 | 307.9 | 2.8 | 0.2 | accelerating decline |
| -3.83 | | | | | | 4 | 323.0 | 17.9 | 0.0 | no change |
| **(D) Measles** | | | | | | | | | | |
| **intercept** | **year** | **year2** | **NA** | **NA** | **NA** | **df** | **AICc** | **dAICc** | **weight** | **shape** |
| -0.78 | -0.90 | -0.96 | | | | 6 | 677.7 | 0 | 1.0 | downward opening |
| -1.76 | -0.86 | | | | | 4 | 685.3 | 7.6 | 0.0 | decline |
| -1.78 | | | | | | 4 | 691.8 | 14.1 | 0.0 | no change |

*Table S6 Changes in parish-specific fade-out probabilities over time are (A) infection-specific with (B) for smallpox a parabolic quadratic association while (C) pertussis and (D) measles show a declining probability. Shape of the associations is indicated by the year coefficients. Shape of the associations is indicated by the year coefficients and are consistent with those parish-specific fade-outs results described in table S3. Model specifications and abbreviations as in Table S3.*

| **Terms** | | | | | | **Model Fit** | | | | **Shape** |
|---|---|---|---|---|---|---|---|---|---|---|
| **(A) Three infections** | | | | | | | | | | |
| **intercept** | **year** | **year2** | **inf** | **year*inf** | **year2*inf** | **df** | **AICc** | **dAICc** | **weight** | |
| 4.52 | -0.28 | -0.24 | + | + | + | 13 | 131783 | 0 | 1.0 | NA |
| 4.24 | -0.28 | 0.02 | + | + | | 11 | 132179 | 395.7 | 0.0 | NA |
| 4.27 | -0.28 | | + | + | | 10 | 132178 | 394.7 | 0.0 | NA |
| 4.21 | -0.05 | 0.00 | + | | | 9 | 132924 | 1140.9 | 0.0 | NA |
| 4.21 | -0.05 | | + | | | 8 | 132922 | 1138.9 | 0.0 | NA |
| 4.20 | | | + | | | 7 | 132924 | 1140.5 | 0.0 | NA |
| 3.66 | | | | | | 5 | 134775 | 2992.3 | 0.0 | NA |
| **(B) Smallpox** | | | | | | | | | | |
| **intercept** | **year** | **year2** | **NA** | **NA** | **NA** | **df** | **AICc** | **dAICc** | **weight** | **shape** |
| 3.43 | 0.33 | 0.19 | | | | 7 | 47738 | 0 | 1.0 | upward opening |
| 3.61 | 0.27 | | | | | 6 | 47838 | 100.3 | 0.0 | increase |
| 3.62 | | | | | | 5 | 48038 | 300.5 | 0.0 | no change |
| **(C) Pertussis** | | | | | | | | | | |
| **intercept** | **year** | **year2** | **NA** | **NA** | **NA** | **df** | **AICc** | **dAICc** | **weight** | **shape** |
| 4.02 | -0.27 | | | | | 6 | 47959 | 0 | 0.7 | decline |
| 4.02 | -0.27 | 0.02 | | | | 7 | 47961 | 1.8 | 0.3 | accelerating decline |
| 4.02 | | | | | | 5 | 47993 | 33.8 | 0.0 | no change |
| **(D) Measles** | | | | | | | | | | |
| **intercept** | **year** | **year2** | **NA** | **NA** | **NA** | **df** | **AICc** | **dAICc** | **weight** | **shape** |
| 4.69 | -0.27 | -0.23 | | | | 7 | 32584 | 0 | 1.0 | downward opening |
| 4.39 | -0.24 | | | | | 6 | 32663 | 79.4 | 0.0 | decline |
| 4.37 | | | | | | 5 | 32704 | 120.0 | 0.0 | no change |

*Fig. S7 Proportion of weeks with fade-outs of smallpox (blue lines) increased within 10 years after the introduction of vaccination, while those of pertussis (green) and measles (red) decreased with time. (A) Best fitting GAM on nationwide fade-outs and (B) the GAM derivative for smallpox showing a significant increase in nation-wide fade-outs starting from 1810 onwards. These results are close to (C) the best fitting GAM for parish-specific fade-outs for which (D) smallpox' GAM derivative show a significant increase in fade-out weeks starting from 1803 onwards. Shaded areas show 95CI.*

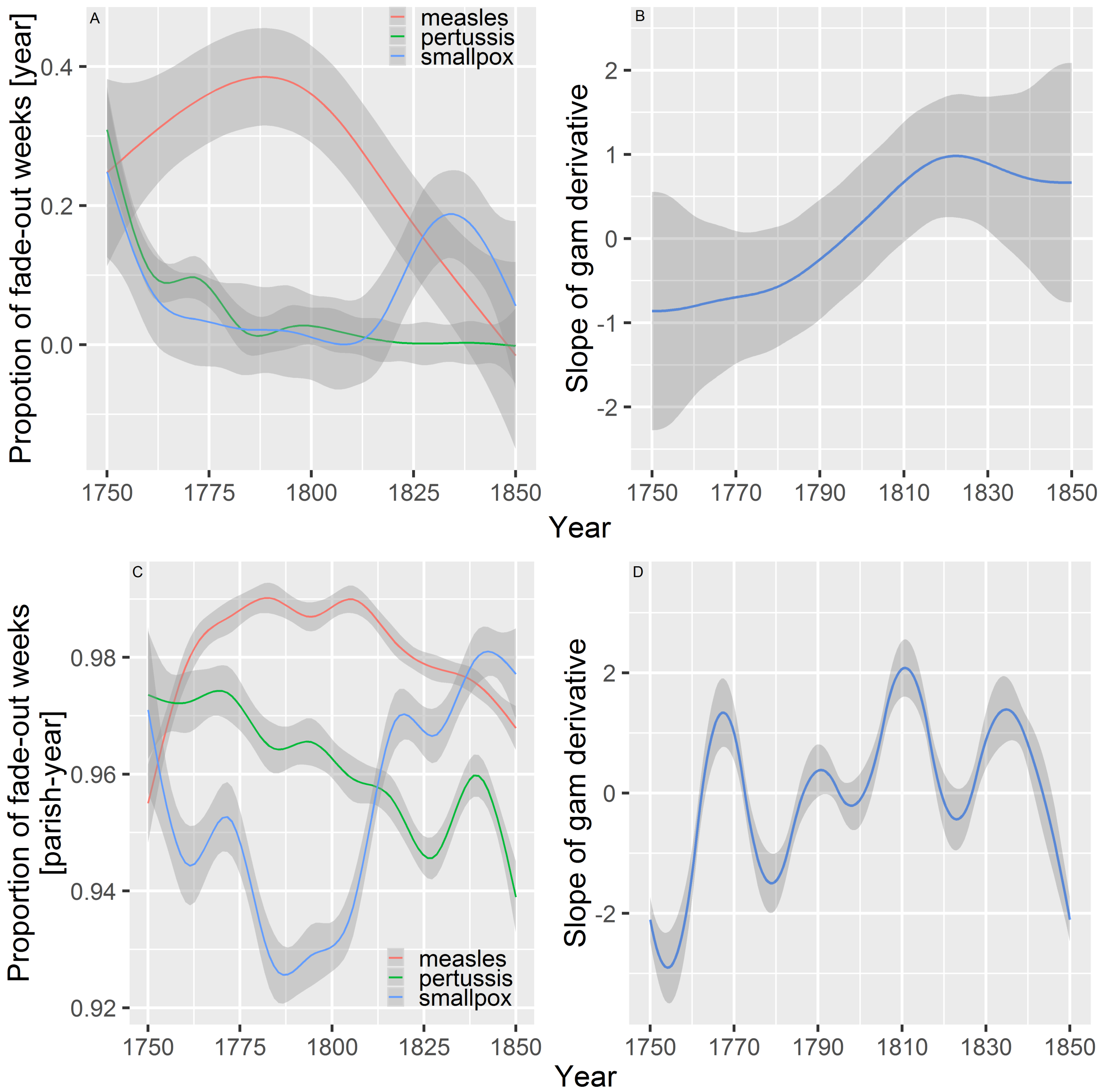

**Supplementary Information 5: Reproduction numbers**

Reproduction numbers $R_0$ can be estimated in many ways and we here used two complementary approaches (Keeling and Rohani 2011; Guerra et al. 2017). First, we used $R_0 \approx \mu^{-1}/A$ where μ is the per capita birth rate and A the average age of infection (Keeling and Rohani 2011). Per capita birth rate in the pre-vaccine era was 0.042 (SD: 0.0027) and decreased in the vaccine era to 0.037 (SD: 0.0032). Second, we estimated $R_0$ using the time-series susceptible-infected-recovered (TSIR) model (Bjørnstad et al. 2002). In brief, in TSIR models the population is divided into Susceptibles S, Infected I and Recovered R. In our study, S was based on the number of births and I on the number of deaths. In this model, $R_0 \approx \beta/\gamma$ in which β captures the rate at which individuals transfer from S to I and is γ the pathogen's generation time (Keeling and Rohani 2011).

Second, to confirm the effect of vaccination on $R_0$ found with mean ages at death, we here tested whether we could detect a change in $R_0$ between the pre-vaccine and the vaccine eras for each infection using the TSIR model. The TSIR model divides the time series in discrete steps which reflects the pathogen's generation time and is two weeks for smallpox and measles and four weeks for pertussis (Keeling and Rohani 2011). To fit the TSIR model on the data, we used the implementation with the function 'estpars' in the R package 'tsiR' (Becker and Grenfell 2017). The first step of the model is to reconstruct the susceptible dynamics. At each time step, the number of susceptibles $S_t$ fluctuates around a mean $S_m$ such that $S_t = S_m + Z_t$. We tested a range of $S_m$ values between 0.03 till 0.30 and fixed $S_m$ values for each infection at values that created the best fitting reconstruction of epidemic dynamics (Fig. S8). These were 0.19 for smallpox, 0.10 for pertussis and 0.15 for measles. The conclusions were robust against changes in $S_m$ values. We assumed there were no differences in $S_m$ values between the pre-vaccine and vaccine eras. Changes in $S_m$ values are possible and actually likely, but including these changes, if they were quantifiable, would exacerbate the result and prevent from estimating the impact of other socio-demographic changes on the estimate of $R_0$.

The deviation susceptibles $Z_{t+1}$ at each time step is then given by:

$$Z_{t+1}=Z_t+B_t-\rho_t I_t. \quad (1)$$

In this equation $B_t$ and $I_t$ are the number of births and infected respectively as given by the data. $\rho_t$ is a time varying correction factor for underreporting. However, because our data reports only deaths, $\rho_t$ here is a combined measure of underreporting and fatality rate. It can be estimated using the regression between cumulative births and cumulative observed cases. Given these parameters, the dynamics of the deviation susceptibles $Z_{t+1}$ can be estimated by the residuals of the regression model in (1), which for small populations as in pre-vaccination Finland, is best captured by a Gaussian process (Caudron et al. 2015).

Following the TSIR model, the infection dynamics at each time step can be described by:

$$\log(E[I_{t+1}])=\log(\beta)+ \alpha \log(I_t) +\log (S_m+Z_t) \quad (2)$$

where the expected number of infected individuals $E[I_{t+1}]$ is given by the data. α is a correction factor that accounts for the discretization of a continuous time process and for inhomogeneous population mixing. We here fixed α=0.97, a common value that generates transmission dynamics independently of epidemic size (Bjørnstad et al. 2002; Caudron et al. 2015; Becker and Grenfell 2017). Our results were robust against small variations around the advised α-value. In equation (2) the only remaining unknown parameter is β, which can be used to estimate $R_0$ by $\beta^*N_m$ in which is $N_m$ is the mean population size per era (Bjørnstad et al. 2002). This can be estimated at each time point as the residuals of the regression of $\log(E[I_{t+1}])$ on $\log(I_t)$ with $\log (S_m+Z_t)$ as an offset. The most exact estimation is achieved using a generalized linear model with a Gaussian distribution and log link function, which is what we use here. Testing other distributions gave consistent results (results not shown).

*Fig. S8 Match between the reconstructed epidemics based on the parameters of the TSIR model (red) and the data (blue) for smallpox (A, B), for pertussis (C, D) and measles (E, F) in the pre-vaccine and vaccine eras respectively.*

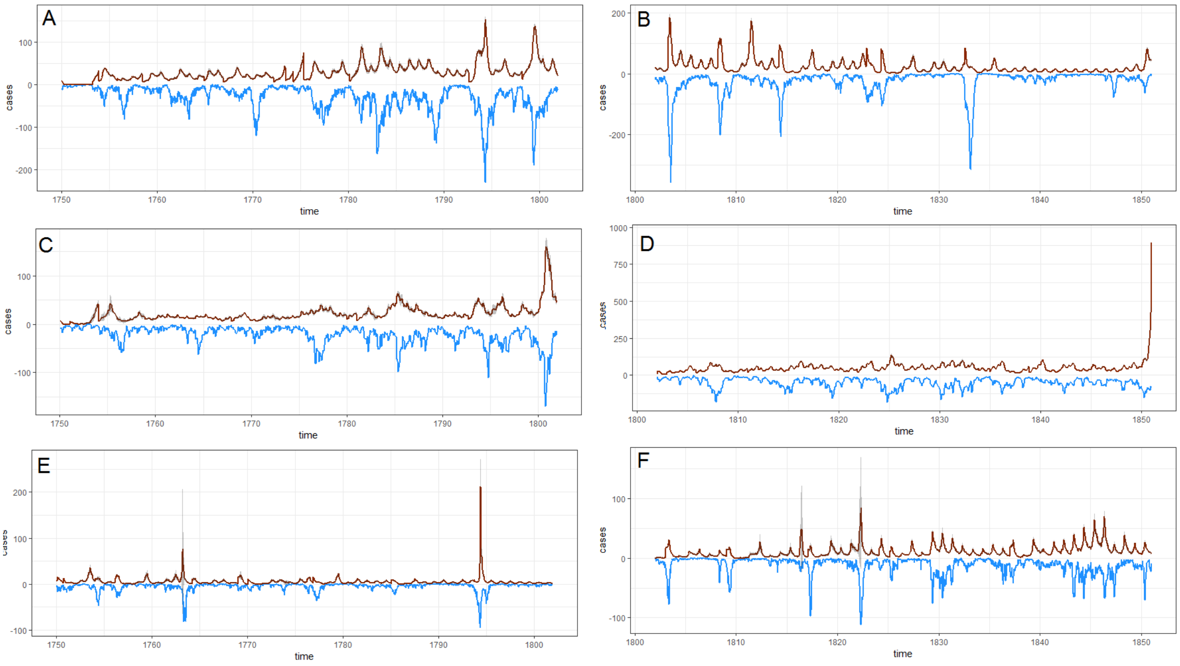
